# Implementation and evaluation of a novel real-time multiplex assay for SARS-CoV-2: In-field learnings from a clinical microbiology laboratory

**DOI:** 10.1101/2020.06.03.20117267

**Authors:** Eloise Williams, Katherine Bond, Brian Chong, Dawn Giltrap, Malcolm Eaton, Peter Kyriakou, Peter Calvert, Bowen Zhang, Mahendra Siwan, Benjamin Howden, Julian Druce, Mike Catton, Deborah A Williamson

## Abstract

The unprecedented scale of testing required to effectively control the coronavirus disease (COVID-19) pandemic has necessitated urgent implementation of rapid testing in clinical microbiology laboratories. To date, there are limited data available on the analytical performance of emerging commercially available assays for severe acute respiratory syndrome coronavirus 2 (SARS-CoV-2) and integration of these assays into laboratory workflows. Here, we performed a prospective validation study of a commercially available assay, the AusDiagnostics Coronavirus Typing (8-well) assay. Respiratory tract samples for SARS-CoV-2 testing were collected between 1^st^ March and 25^th^ March 2020. All positive samples and a random subset of negative samples were sent to a reference laboratory for confirmation. In total, 2,673 samples were analyzed using the Coronavirus Typing assay. The predominant sample type was a combined nasopharyngeal/throat swab (2,640/2,673; 98.8%). Fifty-four patients were positive for SARS-CoV-2 (0.02%) using the Coronavirus Typing assay; 53/54 (98.1%) positive results and 621/621 (100%) negative results were concordant with the reference laboratory. Compared to the reference standard, sensitivity of the Coronavirus Typing assay for SARS-CoV-2 was 100% [95% CI 93.2%-100%], specificity 99.8% [95% CI 99.1%-100%], positive predictive value 98.1% (95% CI 90.2%-99.7%] and negative predictive value 100% [95% CI 99.4%-100%]. In many countries, standard regulatory requirements for the introduction of new assays have been replaced by emergency authorizations and it is critical that laboratories share their post-market validation experiences, as the consequences of widespread introduction of a sub-optimal assay for SARS-CoV-2 are profound. Here, we share our in-field experience, and encourage other laboratories to follow suit.

## 1. INTRODUCTION

The coronavirus disease (COVID-19) pandemic caused by severe acute respiratory syndrome coronavirus 2 (SARS-CoV-2) is a global public health emergency on an unprecedented scale. One of the fundamental pillars in the prevention and control of COVID-19 is timely, scalable and accurate diagnostic testing. Initial laboratory responses included early characterization and release of the viral whole genome sequence by Chinese investigators in early January 2020 ^1^, which enabled rapid development of real-time reverse-transcription polymerase chain reaction (RT-PCR) workflows for detection of SARS-CoV-2. In many settings, testing was initially conducted using in-house RT-PCR assays in public health laboratories ^2-6^. However, the sheer scale of testing required to effectively control this pandemic means there is an urgent need for rapid, sensitive and specific testing in routine clinical microbiology laboratories beyond the public health laboratory setting.

In many countries, the need for rapid introduction of SARS-CoV-2 testing has resulted in rapid changes or extensions to existing regulatory frameworks. For example, in the United States (US) the US Food and Drug Association (FDA) began allowing SARS-CoV-2 testing using laboratory-developed tests without prior agency approval on 29^th^ February 2020, as long as laboratories submitted an Emergency Use Authorization application within 15 days ^7^. In Australia, the Commonwealth Department of Health exempted medical devices related to the diagnosis, confirmatory testing, prevention, monitoring, treatment or alleviation of COVID-19 infection from the requirement for devices to be included in the Therapeutic Goods Administration (TGA) Australian Register of Therapeutic Goods (ARTG) ahead of use in Australia on 31^st^ January, 2020 for laboratories within the Public Health Laboratory Network of Australasia (PHLN) ^8^ and expanded these exemptions to include all accredited pathology laboratories on 22^nd^ March 2020 ^9^.

A range of commercially available RT-PCR assays are now approved for use in diagnostic laboratories in Australia ^10^, although to date, there are little published data on the performance characteristics and implementation of these assays in diagnostic microbiology workflows ^11^. Here, we describe our initial experience using a commercially-available multiplex two-step nested tandem RT-PCR assay for the detection of coronaviruses that infect humans, including SARS-CoV-2. We demonstrate high sensitivity and specificity for this assay, and describe rapid upscaling and integration into our laboratory workflow.

## METHODS

### Study setting, testing timeline and patient populations

This study was conducted in the Department of Microbiology, Royal Melbourne Hospital (RMH). RMH is an academic teaching hospital located in central Melbourne, Victoria, Australia, that has approximately 1,400 beds across hospital and community settings. On 25^th^ January 2020, a dedicated screening clinic for patients with suspected COVID-19 was implemented at RMH ^12^, and on 11^th^ March 2020 a separate clinic was established at RMH for healthcare workers with suspected COVID-19. From 23^rd^ January to 13^th^ March 2020, diagnostic testing of RMH patients for SARS-CoV-2 was performed at the Victorian Infectious Diseases Reference Laboratory (VIDRL). On 3^rd^ March 2020, testing was implemented in the RMH laboratory using the Coronavirus Typing (8-well) panel (AusDiagnostics, Mascot, Australia).

### Patient samples

Study samples were collected as part of routine clinical care between 1^st^ March 2020 and 25^th^ March 2020. Samples comprised combined nasopharyngeal and throat swabs collected in universal transport media (Copan Diagnostics, Murrieta, USA) or Liquid Amies transport media (Copan Diagnostics, Murrieta, USA), sputum, tracheal aspirates, bronchoalveolar lavage and bronchial washings. Victorian Department of Health guidelines during this period limited testing to patients who met at least one clinical (fever or acute respiratory infection) and one epidemiological criteria for COVID-19 (international travel with onset of symptoms within 14 days of return; close contact of confirmed COVID-19 case with onset of symptoms within 14 days of last contact; healthcare or residential aged care workers; aged and residential care residents; patients who are Aboriginal and Torres Strait Islander); or patients admitted to hospital with acute respiratory tract infection and fever ^13^.

### Diagnostic testing

RNA was extracted from 200uL of clinical samples using either the viral RNA mini kit (QIAGEN, Hilden, Germany) on the EZ1 Advanced system (QIAGEN, Hilden, Germany) or the MT-PREP Extractor System (AusDiagnostics, Mascot, Australia). On both platforms, RNA was eluted in 60uL. Extracted RNA was subsequently tested using the Coronavirus Typing assay. This is a two-step, hemi-nested multiplex tandem PCR, with seven coronavirus RNA targets (Table 1) plus a proprietary artificial sequence as an internal control. Currently, it is intended for use as a research use only (RUO) assay, meaning that it can be used for SARS-CoV-2 testing as long as the testing laboratory is able to provide robust validation data. The High-Plex 24 system (AusDiagnostics, Mascot, Australia) was used to perform the two-step real-time RT-PCR. In each run, the following controls were used: (i) a negative control comprising PCR grade sterile water; (ii) a proprietary artificial target sequence used as an internal control to monitor sample inhibition, and (iii) an external positive control, which initially comprised SARS-2-CoV complementary DNA (cDNA), pending the availability of gamma-irradiated SARS-CoV-2 in mid-March.

**Table 1.** Viral targets present in the AusDiagnostics Coronavirus Typing Assay

| Virus assay | Genetic Target of RT-PCR Sequence |
| --- | --- |
| HCoV-HKU1 | Nucleocapsid |
| HCoV-OC43 | Nucleocapsid |
| HCoV-229E | Membrane protein |
| HCoV-NL63 | Membrane protein |
| MERS-CoV | Orf1ab |
| SARS-CoV | Orf1ab |
| SARS-CoV-2 | Orf1ab |

Abbreviations: CoV, coronavirus; HCoV, human coronavirus; MERS, Middle Eastern respiratory syndrome; Orf, open reading frame; SARS, Severe acute respiratory syndrome.

In keeping with national guidelines regarding the validation of new diagnostic assays for SARS-CoV-2 ^14, 15^ and in order to generate sufficient validation data, all samples were initially tested in parallel with Victoria’s virology reference laboratory, VIDRL. At the reference laboratory, testing was first conducted using an in-house assay for the SARS-CoV-2 RdRp gene^16^. If positive, subsequent testing for the SARS-CoV-2 E gene was conducted, using previously published primers ^2^. All samples that tested positive for SARS-CoV-2 using the Coronavirus Typing assay were sent to the reference laboratory for confirmatory testing. Further, a subset of samples that tested negative on the Coronavirus Typing assay were also tested at the reference laboratory in order to provide additional specificity data. Prior to commencement of clinical testing the reference laboratory also provided a blinded quality assessment panel containing a dilution series of samples of standard culture medium spiked in duplicate with gamma-irradiated culture supernatants of a SARS-CoV-2 isolate, described previously ^16^.

Samples that had a negative result on the Coronavirus Typing assay and the VIDRL RdRP assay were considered a true negative result. Samples that had a positive result for SARS-CoV-2 on the Coronavirus Typing assay were tested on both the RdRP and E gene assays at the reference laboratory. Two concordant positive results between the 3 assays were considered a true positive result. Samples with a single positive assay result were considered discrepant.

### Statistical analyses

Sensitivity, specificity, positive predictive value and negative predictive value were reported with 95% confidence intervals. Spearman’s rank correlation was used to examine the association between viral concentration and days from symptom onset. All statistical analyses were performed using R (version 3.53). A p value ≤ 0.05 was considered statistically significant.

## RESULTS

### Characteristics of the study population and diagnostic testing results

Over the study period, a total of 2,673 patient samples were analyzed using the Coronavirus Typing assay. The predominant sample type was a combined nasopharyngeal/throat swab (2640/2673; 98.8%); with lower respiratory tract samples comprising 33/2,673 (1.2%) of samples (18 sputa, 7 tracheal aspirates, 4 bronchial washes and 4 bronchoalveolar lavage specimens). Overall, 1,129 (42%) patients were male. The median age of patients was 35 years [interquartile range (IQR); 28-50 years]. Most samples were collected through the dedicated COVID-19 screening clinic in the Emergency Department (2,002/2,673 samples; 74.9%), with 513/2,673 (19.2%) samples from the dedicated COVID-19 staff testing clinic, 108/2,673 (4%) from other inpatient wards, 30/2,673 (1.1%) from outpatient clinics and 20/2,673 (0.7%) from the intensive care unit.

Of the 2,673 patient samples tested, 54 patients were positive for SARS-CoV-2 (0.02%). Of these, 33/54 were male (61%). The median age of PCR-positive patients was 40 years [IQR 29-51 years]. Forty-one patients (76%) had returned from international travel in the preceding 14 days, 27 patients (50%) were contacts of known cases of COVID-19 and 15 patients (28%) had both of these risk factors. Four patients (7%) were healthcare workers. Four patients (7.4%) required admission to hospital, however no patients required intensive care unit support. One patient had no clear epidemiological link (no international travel or known contact with a COVID- 19 case), suggesting possible community transmission. Of the 54 SARS-CoV-2 positive patients, the median time from onset of symptoms to swab collection was 3 days [IQR; 1-5 days] and the median turn-around time from sample collection to result for SARS-CoV-2 positive results was 13 hours [IQR; 10-21 hours].

### Assay performance

All 54 positive samples were sent to the reference laboratory for confirmation, and a random subset of 621/2,673 negative samples (23%) was also tested at the reference laboratory. Of the 54 positive results, 53 were positive on at least one confirmatory assay at VIDRL. Of the 621 randomly selected negative samples that were sent to the reference laboratory, all were confirmed as negative. Compared to the reference standard, sensitivity of the Coronavirus Typing assay for SARS-CoV-2 was 100% [95% CI 93.2%-100%], specificity 99.8% [95% CI 99.1%-100%], positive predictive value 98.1% (95% CI 90.2%-99.7%] and negative predictive value 100% [95% CI 99.4%-100%].

Importantly, there was a single discrepant sample that tested positive on the AusDiagnostics Coronavirus Typing assay, and negative on the RdRP and E gene assays at VIDRL. This sample had a melt curve at the appropriate temperature on the AusDiagnostics assay and had a low semi-quantitative concentration of SARS- CoV-2. Testing of multiple replicates of this sample at VIDRL disclosed a pattern of results typical of a sample at the limit of detection in the reference assays: positive in some replicates at high cycle threshold values, and negative in others (data not shown). This sample was collected 4 days after symptom onset from a patient with who had recently returned from international travel and had contact with a known case of COVID-19. Given the public health consequences of reporting a false- negative result, we elected to report this result as a ‘probable low positive result.’

The comparative results of the quality assessment panel from the reference laboratory are shown in Supplementary Table 1. All samples were extracted on both the EZ1 Advanced system and the MT-PREP Extractor System. SARS-CoV-2 was detected in all 12 gamma-irradiated SARS-CoV-2 spiked samples from a blinded set of 20 samples using the Coronavirus Typing assay (Supplementary Table 1).

Other respiratory pathogens were detected in 191/2,673 (7%) samples, with seasonal coronaviruses accounting for 119/191 (62%) of these infections. Coinfection with SARS-CoV-2 was detected on two occasions, one in combination with RSV and another with human coronavirus OC43 (Supplementary Table 2).

A weak inverse correlation was demonstrated between the semi-quantitative assessment of viral concentration of SARS-CoV-2 using the Coronavirus Typing assay and the time of swab collection after symptom onset (r^2^ = −0.3357; p=0.01) with the highest viral concentrations detected in the first 24-48 hours after symptom onset (Figure 1).

**Figure 1.**
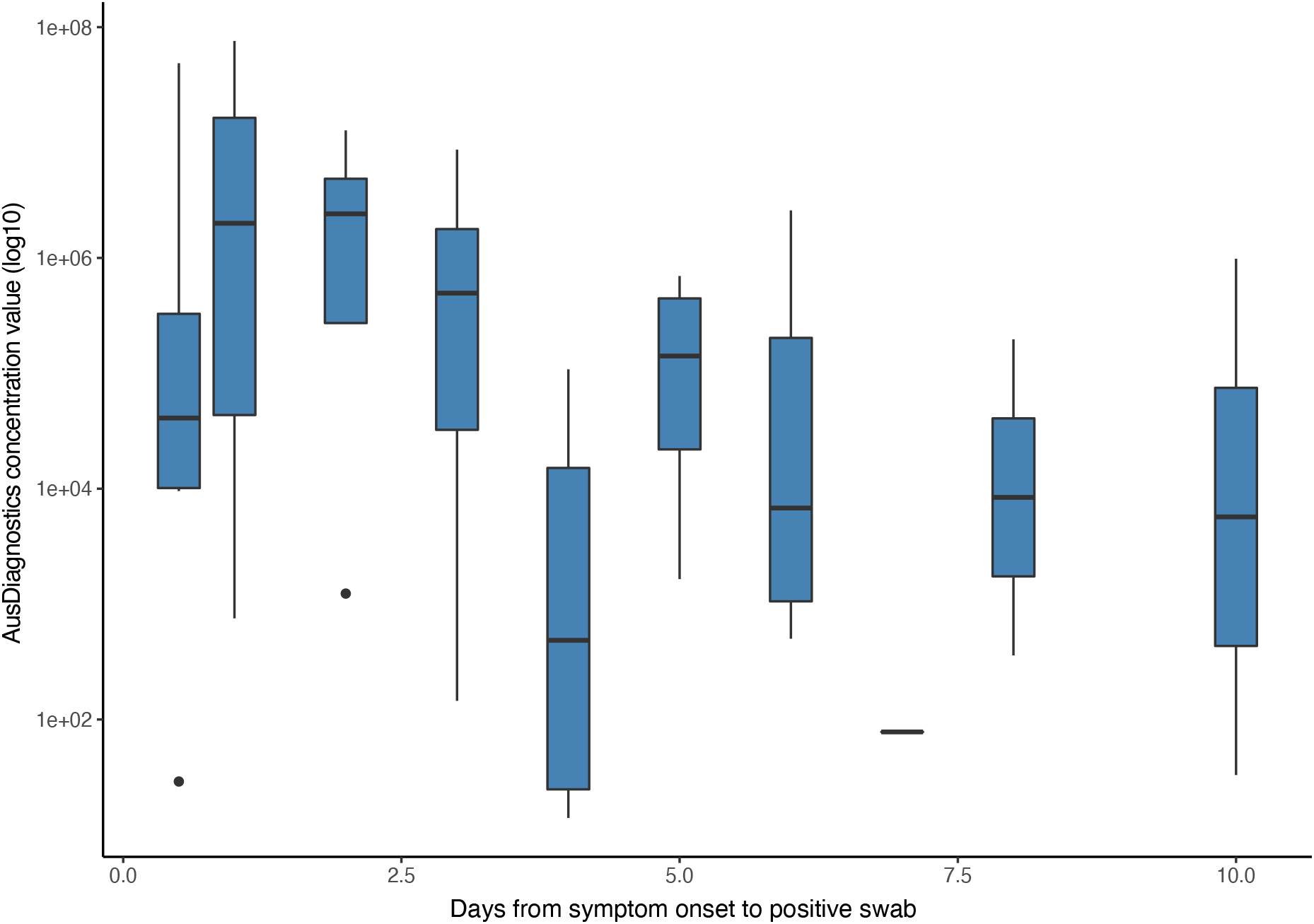
Correlation between semi-quantitative assessment of viral concentration of SARS-CoV-2 and time of swab collection after symptom onset. **Figure 1 Legend**. Box-plot of Coronavirus Typing assay semi-quantitative concentration value relative to the date a nasopharyngeal/throat swab was taken for SARS-CoV-2 after symptom onset. The solid line represent the median and the whiskers represent the interquartile range

### Integration into workflow

The number of samples received for SARS-CoV-2 increased rapidly after the implementation of testing at RMH, with over 200 samples received per day by the second week of testing (Figure 2A). We undertook an internal audit assessing the time of day that specimens arrived in the laboratory; we identified that the majority of samples arrived in the afternoon and evening, rather than in the morning (Figure 2B). This information enabled us to rapidly adjust work rosters for our specialist molecular scientific staff rostering in order to optimize our SARS-CoV-2 testing workflow. This adjustment allowed the laboratory to consistently achieve turnaround times of less than 24 hours for coronavirus testing (Figure 2C).

**Figure 2.**
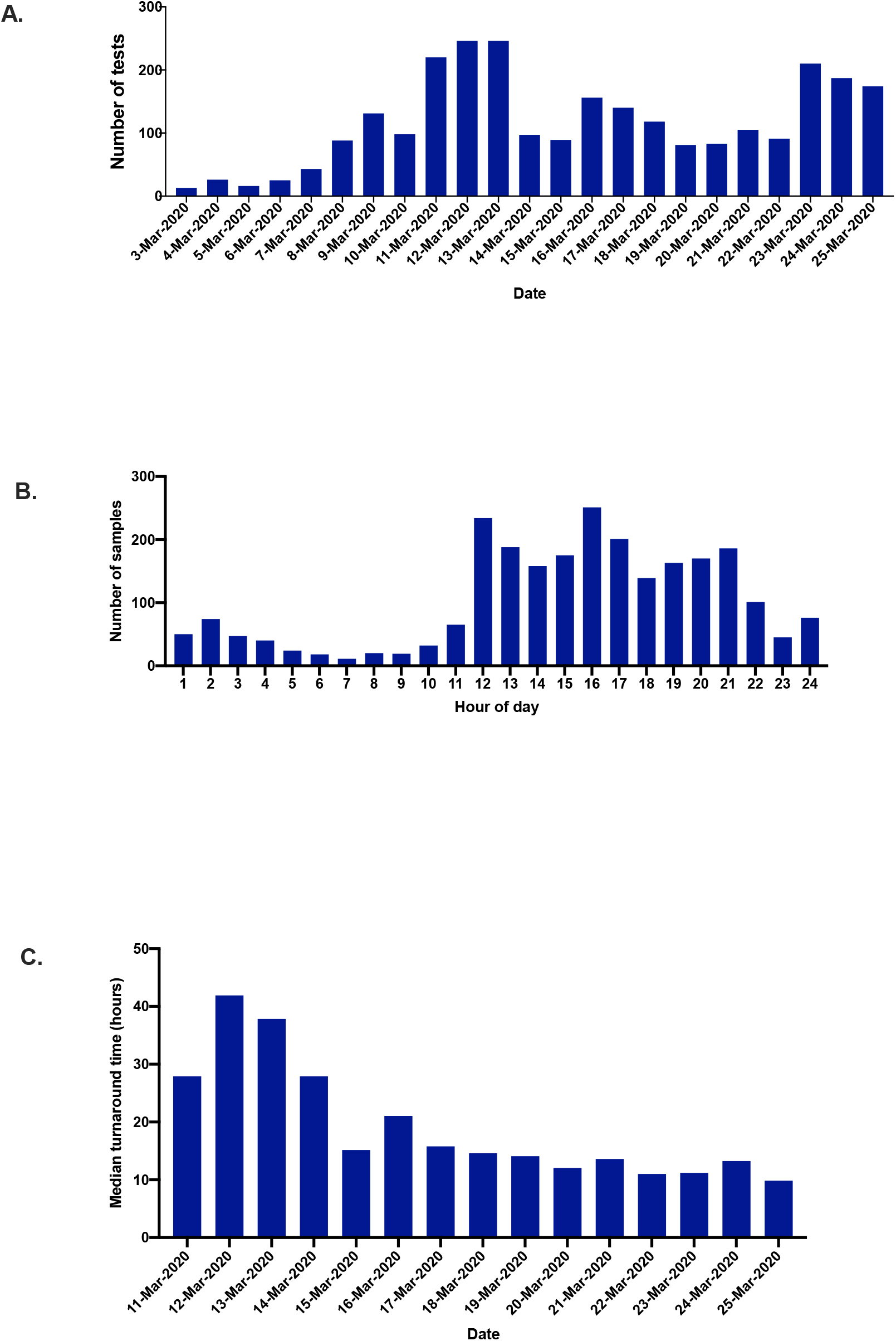
Characteristics of samples received for SARS-CoV-2 testing during the validation study period. **Figure 2 Legend**. A) Number of SARS-CoV-2 tests performed using the Coronavirus Typing assay per day; B) Number of samples received by hour of the day for SARS- CoV-2 during the study period; C) Median turnaround time for SARS-CoV-2 testing from sample collection to result availability per day!

## DISCUSSION

Initial diagnostic testing for SARS-CoV-2 in low prevalence, high resource settings was focused in public health labs, generally using in-house real-time RT-PCR assays recommended to the World Health Organization (WHO) ^2, 15^. However, as the COVID-19 pandemic has rapidly evolved, the focus of testing has shifted to diagnostic testing in clinical microbiology laboratories to enable the delivery of clinical care ^7^. The emergency exemptions instituted by national regulatory bodies such as the FDA in the US and the TGA in Australia have led to expedited approvals for in-vitro diagnostic tests to be generated based only on information provided by the manufacturer and limited external validation (https://www.tga.gov.au/covid-19-test-kits-included-artg-legal-supply-australia). There is thus an urgent need for robust clinical validation data to support the use of commercial assays for SARS-CoV-2 testing. Here, we provide validation of a commercially-available assay, and describe how we integrated testing for SARS-CoV-2 into a busy academic hospital laboratory.

Our early experience of testing for SARS-CoV-2 is similar to other early data from low incidence settings ^17, 18^. This initial cohort of patients with COVID-19 was predominantly returning international travelers; and half had known contact with a confirmed COVID-19 case. In general, patients were younger than those seen in higher incidence settings where community transmission has been established^19^. These findings may be impacted by selection bias, given the Victorian Department of Health guidelines in place at the time of this study mostly limited testing to those with epidemiological risk factors for COVID-19. At the beginning of this study, only limited community transmission had been established in Australia ^20^, with the situation evolving such that on the last date of patient inclusion, 2,799 cases had been confirmed in Australia, with 547 new cases in the previous 24 hours and 11 total deaths ^21^. Significant public health measures designed to mitigate the clinical, societal and economic impact of COVID-19 were instituted in a staged manner during this study period including international travel restrictions and increasing social distancing measures ^22^.

Similar to other studies, we describe a correlation between days after symptom onset and concentration of SARS-CoV-2, with the highest concentrations demonstrated early after symptom onset, ^23-25^. This suggests that SARS-CoV-2 can be transmitted early in disease, even when symptoms are relatively mild and often prior to patients seeking medical attention and the diagnosis of COVID-19 being established. This may account for the efficient person-to-person transmission noted ^26^, particularly within families and social gatherings ^27, 28^. This finding has significant implications for infection control and public health measures required to mitigate this disease.

Compared to a public health laboratory reference standard, we found that the Coronavirus Typing assay had high sensitivity, specificity and negative predictive value, and in our laboratory, was well-suited to medium-throughput testing for SARS-CoV-2 (100-200 specimens per day). We hypothesize that the single discrepant result (positive on Coronavirus Typing assay and negative at reference laboratory) was due to technical differences in the assay design, with the Coronavirus Typing assay having a hemi-nested design and high number of PCR cycles – it is possible that these technical differences may result in a lower limit of detection than the public health laboratory reference standard. In addition, testing on the original referred swab after RMH diagnostic testing had been performed may have diminished the quantity of available input material for testing at the reference laboratory.

In order to accommodate the rapid introduction of a high-impact new test, it was necessary to undertake considerable changes to the workflow of the laboratory, with a change in our staffing shift patterns towards late afternoon and evening work. Similar to many clinical microbiology laboratories, molecular work in our laboratory is performed by staff with specific skills in molecular biology. However, in order to accommodate changes in shift pattern (and to ensure a larger pool of staff to perform testing) we introduced a rapid training program for non-molecular staff. Collectively, these workflow changes enabled us to reduce the turn-around time for producing a SARS-CoV-2 result, which will be important as the pandemic in Australia evolves towards clinical care of infected patients.

Although a limitation of our study is the single-site design, we provide robust validation data for a testing platform that is widely used across Australia, and has also been used in the United States and the United Kingdom ^29^. In this rapidly evolving pandemic, where standard regulatory requirements for the introduction of new assays have been replaced by emergency authorizations in many countries, including Australia, it is critical that laboratories share their post-market validation experiences, as the consequences of widespread introduction of a sub-optimal assay for SARS-CoV-2 are profound.

COVID-19 has placed unprecedented demands on clinical microbiology laboratories. In conjunction with the public health units, clinical microbiology laboratories were part of the first ‘wave’ of response to this pandemic ^30^. Subsequent waves will relate to clinical management, critical care and end of life support for those affected by this disease ^31-33^. In Australia, containment and preparedness measures, particularly international travel restrictions, allowed time for clinical microbiology laboratories to review diagnostic assays, develop workflows and implement testing prior to the surge in demand. Here we share our pragmatic ‘in-field’ experience, and encourage other laboratories to follow suit.

## Data Availability

The datasets generated and analysed during the current study are available from the corresponding author on reasonable request.

## ACKNOWLEDGEMENTS

We thank staff in the Department of Microbiology at the Royal Melbourne Hospital and in the Victorian Infectious Diseases Reference Laboratory for technical support.

## ETHICS

This study was approved as part of routine activities relating to the introduction and validation of in-vitro diagnostic devices QA2019134. Validation studies are a core part of laboratory work. This validation study was conducted according to the guidelines of the National Pathology Accreditation Advisory Council, *‘Requirement for quality control, external quality assurance and method evaluation (sixth edition)’^34^*.

All authors had full access to all of the data (including statistical reports and tables) in the study.

